# Integrating NHANES and Toxicity Forecaster Data to Compare Pesticide Exposure and Bioactivity by Farmwork History and US Citizenship

**DOI:** 10.1101/2023.01.24.23284967

**Authors:** Chanese A. Forté, Jess A. Millar, Justin Colacino

**Author notes:** **Corresponding Author:** Justin Colacino, 6611D SPH1, 1415 Washington Heights, Ann Arbor, MI, 48104.

## Abstract

**Introduction:** Farmworkers in the United States, especially migrant workers, face unique barriers to healthcare and have documented disparities in health outcomes. Exposure to pesticides, especially those persistent in the environment, may contribute to these health disparities.

**Methods:** We queried the National Health and Nutrition Examination Study (NHANES) from 1999-2014 for pesticide exposure biomarker concentrations among farmworkers and non-farmworkers by citizenship status. We combined this with toxicity assay data from the US Environmental Protection Agency’s (EPA’s) Toxicity Forecast Dashboard (ToxCast). We estimated adverse biological effects that occur across a range of human population-relevant pesticide doses.

**Results:** In total, there were 1,137 people with any farmwork history and 20,205 non-farmworkers. Of the 14 commonly detectable pesticide biomarkers in NHANES, 2,4-dichlorophenol (OR= 4.32, p= 2.01×10^−7^) was significantly higher in farmworkers than non-farmworkers. Farmworkers were 1.37 times more likely to have a bioactive pesticide biomarker measurement in comparison to non-farmworkers (adjusted OR=1.37, 95% CI: 1.10, 1.71). Within farmworkers only, those without U.S. citizenships were 1.31 times more likely to have bioactive pesticide biomarker concentrations compared those with U.S. citizenship (adjusted OR 1.31, 95% CI: 0.75, 2.30). Additionally, non-citizen farmworkers were significantly more exposed to bioactive levels of *β*-hexachlorocyclohexane (BHC) (OR= 8.50, p= 1.23×10^−9^), p,p-DDE (OR= 2.98, p= 3.11×10^−3^), and p,p’-DDT (OR= 10.78, p= 8.70×10^−4^).

**Discussion:** These results highlight pesticide exposure disparities in farmworkers, particularly those without U.S. citizenship. Many of these exposures are occurring at doses which are bioactive in toxicological assays.

## 1.1 Introduction

Pesticide exposure has been linked to a myriad of human health outcomes such as obesity, immune alteration, cancer, neurological conditions, type II diabetes mellitus, and death (Wei et al. 2014; Zong et al. 2018; Medehouenou et al. 2019). More specifically, many pesticides are strong endocrine disruptors because they mimic hormones like estrogens and androgens (Briz et al. 2011; Wong et al. 2019). Persistent pesticides last in the environment and human body for years or even decades and can bioaccumulate and bioconcentrate. Persistent pesticides include organochlorines like dichlorodiphenyltrichlorethane (DDT), Lindane, Chlordane, Dieldrin, Heptachlor and their metabolites. Non-persistent pesticides include organophosphates, carbamates, pyrethroids, chlorinated phenols, acyl alanine fungicides and more chemical groups, and were thought to be the less harmful answer to previously used persistent chemicals (e.g. organochlorines) (Abubakar et al. 2020). However, non-persistent chemicals still affect human health. While pesticides are associated with endocrine disruption, cancers, and motor neuron disorders, there is still a lack of human health data on the dose-response, toxicological mechanisms, or how population exposure concentrations relate to social determinants of health (Mostafalou and Abdollahi 2013; Dhananjayan and Ravichandran 2018).

Social determinants of health like occupation or citizenship can alter both exposure and health outcomes related to chemicals like pesticides. Healthcare policy and services are limited to non-existent for immigrants and especially migrant workers residing in the United States (US). For example, many policies that on the surface appear highly beneficial for the American people like the Affordable Care Act of 2010, actually exclude immigrants completely from accessing care (Quesada et al. 2011). In addition, agreements like the North American Free Trade Agreement between the US, Canada, and Mexico limit migrant worker rights (Barnes 2013). Moreover, migrant worker health is often unprotected by the law and workplace discrimination leaves migrant workers very vulnerable (Quesada et al. 2011; Ramos et al. 2016; Ramos 2018; Saxton and Stuesse 2018). Prior research on migrant workers in the US Midwest found factors like economics, logistics, and health significantly affected the mental health of migrant workers (Ramos et al. 2015). Overall, a gap exists in the quantification of pesticide exposure among farmworkers and migrant workers, and specifically how these exposures may differ by worker category or US citizenship status.

A major challenge in the field of occupational and environmental health is understanding and predicting the health effects of exposure to chemicals like pesticides. There are currently 85,000 chemicals on the global market that Toxic Substances Control Act (TSCA) has listed in its inventory of substances, and there is little to no experimental toxicology or epidemiology data on many of them (Attene-Ramos et al. 2013; Adeola 2021). In 2008, the US Environmental Protection Agency (EPA) collaborated with multiple other federal agencies including the Food and Drug Administration and the National Institute of Environmental Health Sciences to create the Toxicology in the 21^st^ Century (Tox21) program (Thomas et al. 2018). The goal of Tox21 is to develop high throughput testing methods to determine the safety of chemicals such as food additives and pesticides. Additionally, Tox21 quantifies the biological mechanisms that chemicals alter to prioritize the chemicals being tested and generate a wealth of data to predict toxicological responses in the human body (Attene-Ramos et al. 2013; Thomas et al. 2018). These data are a rich, but untapped, resource to characterize the dose-dependent effects of exposure to pesticides in the context of social determinants of health like occupation and citizenship. This data is then presented in the Toxicity Forecast Dashboard (ToxCast).

To address these gaps and understand how pesticide exposure and effects vary by occupation and citizenship, this study’s goal is to determine if people residing in the US are exposed to bioactive concentrations of pesticides. This project has the following aims: 1) quantify and compare pesticide biomarkers among farmworkers and non-farmworkers, 2) quantify and compare pesticide biomarkers between citizen and non-citizen farmworkers, 3) compare exposure concentrations to known bioactive benchmark concentrations in the Tox21 high throughput toxicity data (ToxCast). We hypothesized that on average farmworkers will have higher concentrations of pesticides biomarkers than non-farmworkers. Furthermore, among farmworkers, we hypothesize that non-citizens will have higher pesticide biomarker concentrations than US citizens. Additionally, we hypothesize people residing in the US will be exposed to bioactive concentrations of pesticides. Moreover, we hypothesize farmworkers will be exposed to bioactive concentrations of pesticides more frequently than non-farmworkers.

## 1.2 Methods

Our overall study design involves comparing the distributions of chemical biomarker concentrations in The National Health and Nutrition Examination Survey (NHANES) with the distributions of doses for those chemicals which exhibit bioactivity in ToxCast. In addition, we quantify which cellular target families are most often affected by these pesticides and look to see how these target families differ by history of farmwork and U.S. citizenship status.

### 1.2.1 The National Health and Nutrition Examination Survey (NHANES)

NHANES is a cross-sectional study representative of the US population with oversampling weights for minoritized populations. NHANES is a cross sectional assessment of the health and nutrition of adults and children residing within the US. The current iteration of the continuous study began in 1999. Study participants are enrolled on a continuous basis, with data analyzed and deposited in two-year windows. NHANES collects extensive information on the study participants such as self-reported occupation, urinary and serum biomarkers, and self-reported demographics such as age, gender, citizenship, poverty index ratio, and education.

### 1.2.2 Study Population

This study included NHANES study participants aged 18 years and older who also had occupation and pesticide exposure data present between 1999 and 2014. This study integrated 29 datasets from NHANES laboratory data to understand pesticide exposure, occupation, and demographics of the study population. From the Industry and Occupation Survey, individuals were coded as “farmworker” or “non-farmworker” using the Current Industry (OCD230=1, OCD231=1), Current Occupation (OCD240=18, OCD241=18), Longest Industry (OCD390=1, OCD391=1), and Longest Occupation (OCD392=18), where all participants who put “Agriculture, Forestry and Fishing” were coded as a farmworker.

From the demographics data, DMDEDUC2 (older than 18 years of age) and DMDEDUC3 (l8 years of age and younger) were combined to create one education level based on the DMDEDUC2 categories. The US citizenship variable (DMDCITZN) is defined as 1= “Citizen by Birth or naturalization” and 2= “Not a citizen of the US”, and we removed anyone who responded with “Refused”, “Don’t Know”, or skipped the question.

### 1.2.3 Biomonitoring Samples and Detectability

NHANES performs chemical biomonitoring in study participants urine and blood. Participants provided partial urine void in a sterile sampling cup at the mobile examination center. Blood samples are collected by certified laboratory professionals. Urine and blood samples are then analyzed for chemical metabolites using isotope dilution gas chromatography high-resolution mass spectrometry (GC/IDHRMS). Pesticide biomarkers measured in blood samples and reported as either 1) fresh weight basis (i.e., pg/g serum) and 2) lipid weight basis (i.e., ng/g lipid). The lipid adjusted values account for blood lipid concentrations and are of particular importance for the accurate quantification of lipophilic pesticides (Barr et al. 2005).

All urinary biomarker measurements were adjusted for urinary creatinine, and all blood pesticide biomarker measurements were blood lipid adjusted. Detectability percentages were calculated by dividing the total number of measurements above LOD by the total number of the chemical’s measurements in NHANES. To ensure that we included chemicals with values above the limit of detection in most of the study participants, detection frequency percentages of 50% and higher across the population were maintained which resulted in 14 chemicals of interest (Silver et al. 2018).

These chemicals included the following: 2,4-Dichlorophenol (24DCP), 2,4-Dichlorophenoxyacetic acid (24D acid), 2,5-Dichlorophenol (25DCP), 3,5,6-Trichloropyridinol (TCP), 4-Nitrophenol, β-hexachlorocyclohexane (β-HCH), diethyltoluamide acid (DEET acid), Dieldrin, Heptachlor Epoxide, 3-phenoxybenzoic acid (3-PBA), p,p’-DDE, and p,p’-DDT. Additionally, the measurements of TCP, a chlorpyrifos metabolite, were compared to the ToxCast toxicity data for both CPF and chlorpyrifos-oxon (CPO).

### 1.2.4 Toxicity Forecast Dashboard Data

The US EPA’s Toxicity Forecast Dashboard (ToxCast) is a collection of publicly available high throughput toxicity data intended to make chemical assessment more accessible by allowing researchers to search which chemicals show toxicological effects more easily within human tissue. High throughput toxicity screening initiatives have been developed to quantify biological effects of chemicals, including pesticides, *in vitro*. Dose response curves are created for each chemical and assay, and from these curves the activation concentrations and positive hitcalls are defined. ACC is the concentration at which the model reaches the cut-off values for the chemical to be considered active and is based on the levels of significance for the dose curve response. The ACC can be used as a proxy of potency to determine the genes, proteins, enzymes, effects on biological pathway and viabilities at which chemicals are active.

### 1.2.5 Comparing NHANES and ToxCast

Using the corresponding Chemical Abstracts Service Registry Numbers (CASRNs) obtained from PubChem, data from ToxCast were matched to NHANES. From this new dataset, we created pesticide concentration distribution boxplots by the chemical and farmwork history or U.S. citizenship in the *tidyverse* using the *ggplot2* R package (Wickham 2016). Pesticide distributions were overlaid unto the same axis to quantify overlap between the pesticide concentration distributions of exposure in NHANES participants and bioactivity in ToxCast. To visualize the distribution of exposure in comparison to pesticide bioactivity concentrations, ToxCast ACCs and NHANES biomarker concentrations were plotted as boxplots using molarity units.

### 1.2.6 Statistical Analysis

All data management and analysis were completed in R version 4.1.3. All code for our work can be found on our GitHub repository (Millar and Forté 2023). Graphics were created using the *ggplot2* package library (Wickham 2016). All NHANES data was downloaded using the RNHANES packaged in R (Susmann 2016). The main outcomes of this project include 1) quantifying the distribution of the pesticide concentrations across NHANES and ToxCast, 2) quantifying the demographics of people with and without bioactive measurements, and 3) investigating how bioactivity differs by chemical, farmwork history, and US citizenship status. These outcomes inform the overarching project question of whether people residing in the US are exposed to bioactive levels of pesticides, how these bioactive pesticides affect the body, and whether the rates of exposure to bioactive pesticide concentrations vary based on sociodemographic factors.

We labeled anyone who had at least one chemical measurement equal to or above the minimum ToxCast ACC for that chemical as being “bioactive”. Anyone who did not fit this group was defined as “non-bioactive.” Demographics were quantified by bioactivity status among all study participants and then among farmworkers only. For continuous variables like body mass index (BMI) or age in years, we present the mean and standard error, and for all categorical variables, the stratified frequencies and sub-group percentages are provided.

Differences in demographic factors by group or citizenship were tested using a Pearson’s chi-square test, using a Rao and Scott Adjustment where necessary for categorical variables. Low response was defined as 8 or less respondents within one stratum. And for continuous variables, a Wilcoxon Rank test was used to test group means, with a Kruskall-Wallis Correction. All significance testing was completed using the NHANES Full Sample 2 and 4 Year MEC Exam Weights. A new weight variable titled “MEC16YR” was created using the weighted MEC 2- and 4-year measurements to represent the weights used from 1999-2002 and each year after, respectively.

Non-citizen status was determined by the NHANES variable DMDCITZN. We calculated bioactivity by the chemical and marked measurements as bioactive based on their hitcall equaling 1. For model outcomes this bioactivity status by chemical was used as the outcome variable for logistic regression models used to investigate how the odds of being a farmworker and having at least one bioactive measurement differ from non-farmworkers by the chemical. These models were adjusted for BMI, age, poverty index ratio (PIR), survey year, gender, racial ethnicity, U.S. citizenship status, farmwork history, country of birth and education level. After comparing all study respondents’ odds of having a bioactive measure, we created logistic regression models comparing U.S. citizenship status. These models were also adjusted for BMI, age, PIR, survey year, gender, racial ethnicity, country of birth, and education level.

Education status was constructed NHANES variables DMDEDUC2 and DMDEDUC3 to include four categories: Less than 9th grade, 9-11th grade (Includes 12th grade with no diploma), High school grad/GED or equivalent, and More than high school. Farmworker status was constructed using NHANES industry or occupation group codes for current job (OCD230, OCD231) or longest job (OCD390, OCD391, OCD392) that included the terms agriculture/agricultural or farming.

For lipid adjusted blood measurements, molarity was calculated by multiplying the measurement by serum density of 1.024 g/mL and dividing by molecular weight (Sniegoski and Moody 1979). Urinary measurements were calculated by diving the measurement by molecular weight. All measurements of molarity have units of μmol/L.

Data from the 1999-2002, 2003-2004, 2005–2006, 2007–2008, 2009–2010, 2011–2012 and 2013-2014 data collection cycles were appended, and the sampling weights modified as directed in NHANES documentation. Removal of observations with missing data was done for all analyses. Statistical analysis was done with the R *survey* package (v4.1-1) to handle complex survey designs present in NHANES. The function *survey::svydesign* was used to handle sampling weights, with primary sampling units nested within each stratum.

Wilcoxon Mann Whitney U test was conducted on individual chemicals in relation to farmworker or non-citizen status using the *survey::svyranktest* function. The outcome variable for chemicals was calculated as the log molarity for blood measurements and the log of the ratio of the chemical molarity to creatine molarity for urinary measurements. P-values for all tested chemicals were FDR adjusted and AUCs were calculated using the U statistic (Mason and Graham 2002).

Both unadjusted and adjusted logistic regression was conducted on individual chemicals in relation to farmworker or non-citizen status using the *survey::svyglm* function using a quasi-binomial model with a logit link. The outcome variable for chemicals was constructed as an indicator variable, with a 1 indicating the measurement was considered chemically bioactive. Adjusted logistic regression included variables for age at screening, race-ethnicity, BMI, education, and survey year for all chemicals, and the additional inclusion of creatine molarity for urinary measurements. P-values for all tested chemicals were FDR adjusted and AUCs were calculated using the *WeightedROC* R package (v2020.1.31) (Hocking 2020).

Initially, the list of pesticides under investigation included 96 different biomarkers present in NHANES, but after removing chemicals with detectability percentages below 50%, we were left with 16 chemicals for analysis (Supplementary Table 1). Assay data for these chemicals from NHANES were then extracted from the ToxCast database. We retrieved the hitcall (representative of an active assay), the activity concentration at cutoff (or ACC), and the intended target family of each ToxCast assay based on the 16 pesticides from NHANES. Using the hitcall variable, we labeled assays as positive (hitcall==1) or negative (hitcall==0) to mean that an assay did or did not show bioactivity by the pesticide. We created a bioactivity ratio per chemical by dividing the number of positive assays by total number of assays. All chemicals in NHANES were present in ToxCast. However, trans-nonachlor was not maintained in the study because there were only 8 completed assays in ToxCast and none of those assays were active.

## 1.3 Results

We first assessed demographic features of the study participants based on whether the participant had a history of farmwork or not (Tables 1 and 2). In total, there were 1,137 people who reported any farmwork history, and 20,205 who were categorized as non-farmworkers. The farmworker group was mostly women (N=697, 61.3%), Non-Hispanic White (N=635, 55.8%), U.S. Citizens (N=934, 82.1%) and 26.6% reported some college education or an associate’s degree (N=302). The non-farmworker group had similar mean BMI, age, and poverty index ratio. The non-farmworker group is predominantly men (N=10,187, 50.4%), Non-Hispanic White (N=9,167, 45.4%), had U.S. Citizenship (N=17,626, 87.2%), and 19.2% reported some college or an associate’s degree (N=3,885).

**Table 1.**
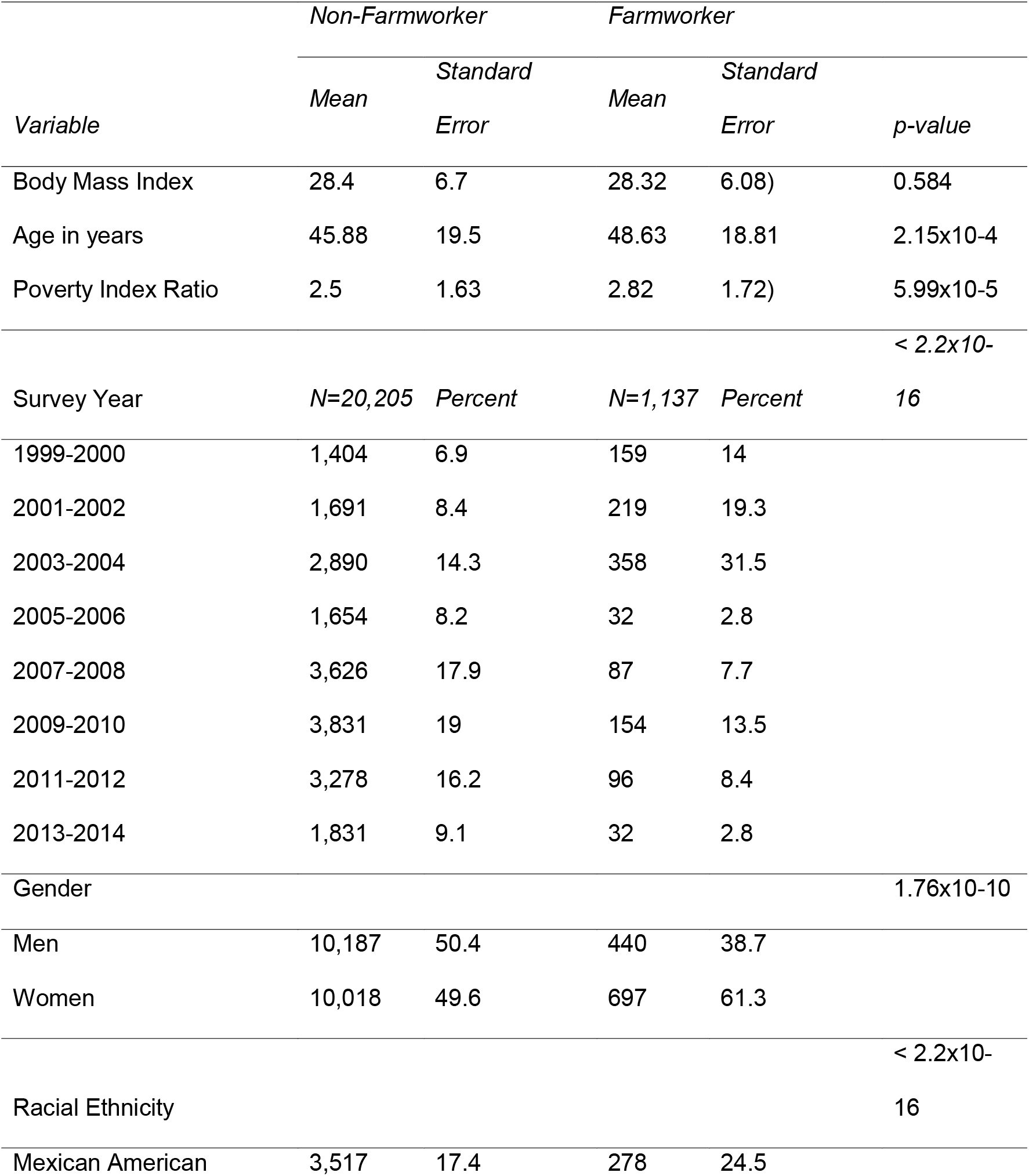
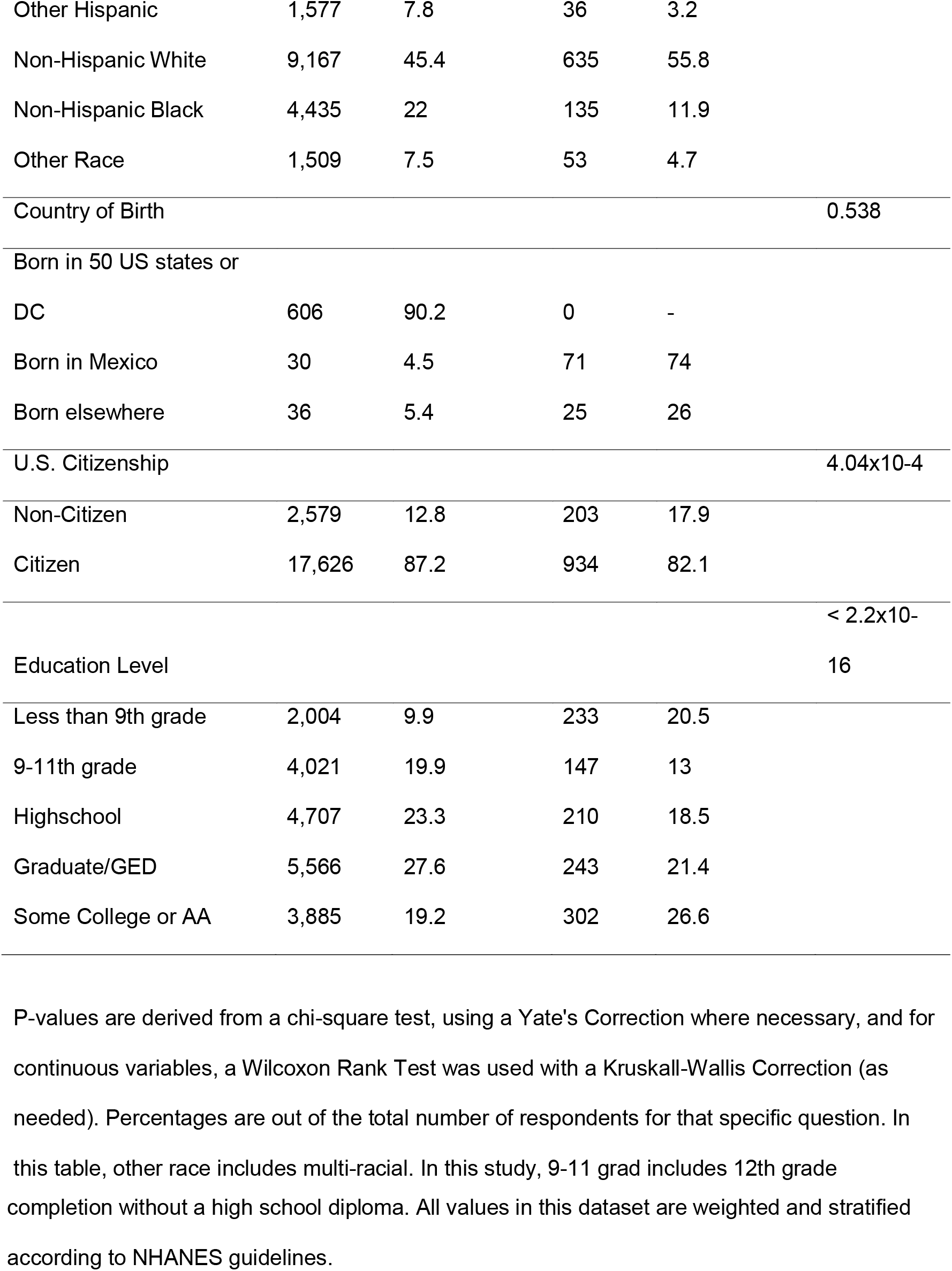
Stratified Demographics of NHANES Participants, by Farmwork Category

**Table 2.** Stratified Demographics of NHANES Participants with a History of Farmwork, by Citizenship

|  |  | *Citizen* |  | *Non-Citizen* |  | *p-value* |
| --- | --- | --- | --- | --- | --- | --- |
| *Variable* |  | *Mean* | *Standard Error* | *Mean* | *Standard Error* |  |
| *Body Mass Index* |  | 28.52 | 6.33 | 27.37 | 4.66 | 0.038 |
| *Age in years* | | 49.9 | 18.9 | 42.74 | 17.07 | $7.57 \times 10^{-5}$ |
| *Poverty Index Ratio* | | 3.13 | 1.68 | 1.41 | 1.07 | $< 2.2 \times 10^{-16}$ |
| *Variable* |  | N=1,007 | % | N=237 | % |  |
| *Survey Year* | *1999-2000* | 139 | 14.9 | 20 | 9.9 | $9.41 \times 10^{-05}$ |
|  | *2001-2002* | 188 | 20.1 | 31 | 15.3 |  |
|  | *2003-2004* | 325 | 34.8 | 33 | 16.3 |  |
|  | *2005-2006* | 20 | 2.1 | 12 | 5.9 |  |
|  | *2007-2008* | 66 | 7.1 | 21 | 10.3 |  |
|  | *2009-2010* | 105 | 11.2 | 49 | 24.1 |  |
|  | *2011-2012* | 68 | 7.3 | 28 | 13.8 |  |
|  | *2013-2014* | 23 | 2.5 | 9 | 4.4 |  |
| *Gender* | *Men* | 378 | 40.5 | 62 | 30.5 | 0.013 |
|  | *Women* | 556 | 59.5 | 141 | 69.5 |  |
| *Racial Ethnicity* | *Mexican American* | 125 | 13.4 | 153 | 75.4 | $< 2.2 \times 10^{-16}$ |
|  | *Other Hispanic* | 23 | 2.5 | 13 | 6.4 |  |
|  | *Non-Hispanic White* | 622 | 66.6 | 13 | 6.4 |  |
|  | *Non-Hispanic Black* | 129 | 13.8 | 6 | 3 |  |
|  | *Other Race* | 35 | 3.7 | 18 | 8.9 |  |
| *Country of Birth* | *Born in 50 US states or DC* | 606 | 90.2 | 0 | 0 | < 2.2 x10<sup>-16</sup> |
|  | *Born in Mexico* | 30 | 4.5 | 71 | 74 |  |
|  | *Born elsewhere* | 36 | 5.4 | 25 | 26 |  |
| *Education Level* | *Less than 9th grade* | 110 | 11.8 | 123 | 60.6 | < 2.2 x10<sup>-16</sup> |
|  | *9-11th grade* | 115 | 12.3 | 32 | 15.8 |  |
|  | *Highschool* | 183 | 19.6 | 27 | 13.3 |  |
|  | *Graduate/GED* | 234 | 25.1 | 9 | 4.4 |  |
|  | *Some College or AA* | 290 | 31.1 | 12 | 5.9 |  |

P-values are derived from a chi-square test, using a Yate's Correction where necessary, and a Wilcoxon Rank Test was completed with a Kruskal-Wallis Correction. Percentages are out of the total number of respondents for that specific question. In this table, other race includes multi-racial. In this study, 9-11 grad includes 12th grade completion without a high school diploma.

To better understand how each of the chemicals relate to each other, Table 3 outlines the pesticides by persistence and frequencies of activity of ToxCast assays. In total, there are 15 pesticides that are detectable in NHANES study participants and also assayed in ToxCast. Overall, there were 5 persistent organic pesticides and 10 non-persistent pesticides included in this study. The top three most bioactive pesticides in ToxCast were heptachlor epoxide had the highest percentage of assays which were “active” (39.85%), followed by p,p’-DDT (35.73%) and p,p’-DDE (26.78%). The bioactivity threshold is the lowest ACC of the active assays for a given chemical. These values ranged from 6.5nM (2,4-Dichlorophenoxyacetic acid) to 1.45μM (chlorpyrifos).

**Table 3.** Bioactivity of pesticides cross-listed between NHANES and ToxCast, by pesticide and persistence

| *Common Name* | *CAS-RN* | *Total*<br>*Assays* | *Positive*<br>*Assays* | *Bio-active*<br>*Assay*<br>*Percentage* | *Bioactivity*<br>*Threshold (μM)* |
| --- | --- | --- | --- | --- | --- |
| 2,4-Dichlorophenol | 120-83-2 | 678 | 27 | 3.98 | 0.34 |
| 2,4-Dichlorophenoxyacetic acid | 94-75-7 | 807 | 18 | 2.23 | 6.49x10<sup>-3</sup> |
| 2,5-Dichlorophenol | 583-78-8 | 599 | 14 | 2.34 | 0.33 |
| 3-Phenoxybenzoic acid | 3739-38-6 | 622 | 11 | 1.77 | 0.23 |
| 3,5,6-Trichloropyridinol | 6515-38-4 | 433 | 21 | 4.85 | 1.35 |
| 4-Nitrophenol | 100-02-7 | 682 | 43 | 6.30 | 8.63x10<sup>-3</sup> |
| β-hexachlorocyclohexane<br>a | 319-85-7 | 654 | 24 | 3.67 | 0.03 |
| Chlorpyrifos | 2921-88-2 | 639 | 126 | 19.72 | 1.45 |
| Chlorpyrifos-oxon | 5598-15-2 | 693 | 132 | 19.05 | 0.04 |
| DEET Acid | 134-62-3 | 1025 | 13 | 1.27 | 0.17 |
| Dieldrin <sup>a</sup> | 60-57-1 | 549 | 121 | 22.04 | 0.32 |
| p,p'-DDE <sup>a</sup> | 72-55-9 | 1139 | 305 | 26.78 | 0.31 |
| p,p'-DDT <sup>a</sup> | 50-29-3 | 778 | 278 | 35.73 | 0.43 |
| Heptachlor Epoxide <sup>a</sup> | 76-44-8 | 650 | 259 | 39.85 | 1.31 |

<sup>a</sup>Persistent Organic Pollutant.
A positive assay is defined as hitcall==1. The bioactivity assay percentage is created by dividing the total number of positive assays by the total number of assays and multiplying by 100%.
Bioactivity ratio per chemical was calculated by dividing the count of positive assays by the total number of assays within the US Environmental Protection Agency's Toxicity Forecast
Dashboard database.

Next, we wanted to compare the concentrations of chemicals required to activate the ToxCast assays to the biomarker concentrations measured in people in NHANES. Figure 1 presents the distribution of pesticide concentrations among people residing in the United States in orange (retrieved from NHANES), and in blue, the ACCs of active assays retrieved from ToxCast. In this figure, where the pesticide distributions of exposure and bioactivity overlap represents pesticide exposures among the US population that are “bioactive”. Additionally, 4-nitrophenol is the only pesticide biomarker in NHANES that does not have human measurements that overlap with the bioactive distribution in NHANES.

**Figure 1.**
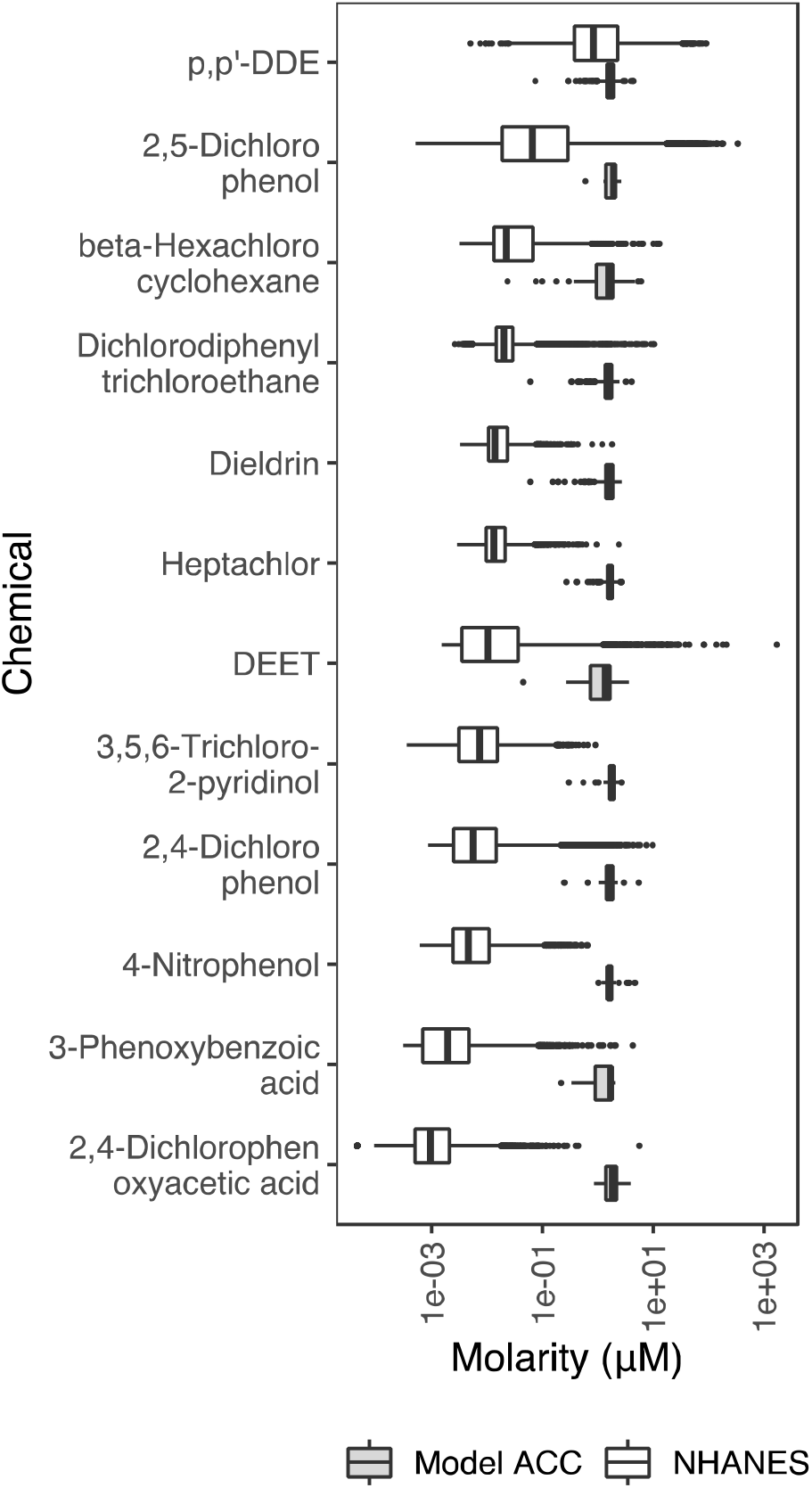
Comparing the chemical molarity of of NHANES subjects with bioactivity threshholds taken from chemical assays. ACC is the activity concentration at cut-off for a specific assay where a chemical is considered active.

We present the Mann-Whitney-U Rank Test outcomes by chemical in Supplementary Table 2 to test for differences in biomarker concentration by farmworker status, or within farmworkers, comparing between farmworkers with and without US citizenship. When quantifying the odds of having a bioactive measurement (unadjusted outcomes in Supplemental Table 3, fully adjusted outcomes presented in Figure 2 and Supplementary Tables 4 and 5), we found farmworkers were 4.3 times more likely to have a bioactive measurement in comparison to non-farmworkers for 2,4-D (p=2.0×10^−7^) while farmworkers were significantly less likely to have a bioactive measurement of 4-Nitrophenol (p= 2.7×10^−4^). Next, we narrowed our analyses to farmworkers only and found farmworkers living without U.S. citizenships were significantly more likely to be exposed to a bioactive measurement of BHC (OR=8.4, p-value=1.2×10^−9^, U=13.95), p,p’-DDE (OR=3.0, p-value=3.1×10^−3^, U=9.43), p,p-DDT (OR=10.8, p-value. =8.7×10^−4^, U=6.56).

**Figure 2.**
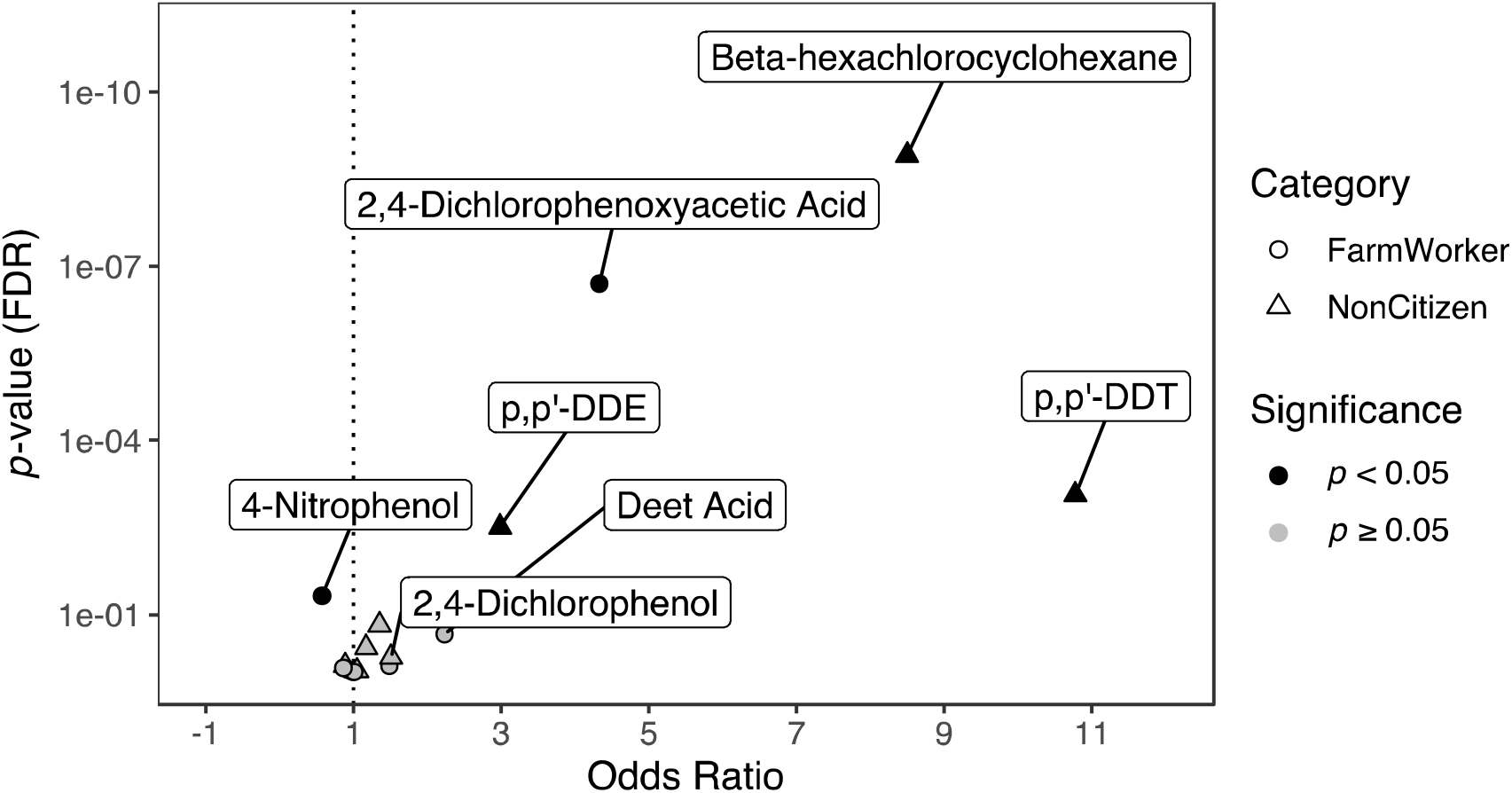
Comparing the odds of having a bioactive pesticide biomarker concentration by farmwork history and for farmworkers only by citizenship. This figure presents the outcomes of the regression model of farmworker and non-farmworker health outcomes. Bioactive was defined as having at least one pesticide biomarker concentration that was the same or higher concentration than the minimal concentration needed to see an effect. The data for this table was retrieved from the U.S. EPA’s Toxicity Forecast Dashboard and the National Health and Nutrition Examination Survey.

When trying to understand what intended target families are most affected by these chemicals, Supplementary Table 6 provides the frequency of intended target families by the pesticide. Based on individual intended assay target count, cell cycle (N=487), nuclear receptor (N=318), cytokine (N=143), DNA binding (N=172), and cell adhesion molecules (N=65) were the most frequent targets of the pesticides. Overall, p,p’-DDE (N=305) had the most intended target family counts based on positive assays, followed by p,p’-DDT (N=278), heptachlor epoxide (N=259), and chlorpyrifos (N=126). Heptachlor epoxide had the highest number of positive assays targeting the cell cycle (N=123) and p,p’-DDT had the second most (N=120). Additionally, for p,p’-DDE had mostly nuclear receptor targeting positive assays (N=102), followed by the cell cycle (N=74) and DNA binding (N=64).

### 1.3.1 Discussion

When looking at individuals who have pesticide biomarker concentrations at these bioactive levels, demographics statistically differed based on bioactivity, farmwork history and citizenship status. We found NHANES participants are broadly exposed to bioactive concentrations of pesticides. Heptachlor epoxide, p,p’-DDT, and p,p’-DDE were the most bioactive pesticides in ToxCast based on overall percent of positive assays. Disproportionate exposures to bioactive concentrations of pesticides were particularly evident in farmworkers without U.S. citizenship, particularly for persistent pesticides.

Pesticide exposures have been associated with increased mortality due to cancer, diabetes mellitus, poisonings, and tuberculosis and other lung infection (Mills et al. 2006; Fry and Power 2017). Pesticide exposure throughout the life course has been associated with breast cancer and dysregulated mammary gland development. For example, mothers with the highest p,p-DDT concentrations were 3.7 times more likely to have daughters who developed cancer by the age of 52 in comparison to mothers with the lowest p,p-DDT blood concentrations (Cohn et al. 2015). Women who are farmworkers and not US citizens could be at increased risk of exposure-associated diseases like breast cancer – these findings warrant further investigation in this area.

Citizenship status is also a known barrier to health insurance and treatment (Guadamuz et al. 2020; Chasens et al. 2020), potentially compounding adverse effects of exposure to toxic chemicals like pesticides. In a study of 2,702 participants living with diabetes, non-citizens had a greater risk for poor glycemic management (OR=5.16, 95% CI: 3.73, 6.04) in comparison to citizens by birth (Chasens et al. 2020). Additionally, citizens by naturalization were also at an increased risk of poor glycemic management (OR=1.95, 95% CI: 1.49,2.55) (Chasens et al. 2020). Additionally, this study found that individuals with diabetes and without health insurance were almost twice as likely to have poor glycemic management compared to insured people (OR=1.99, 95% CI: 1.53-2.59). Similar outcomes have also been noted in cardiovascular disease. Using NHANES, researchers retrieved data from 2011 to 2016 to investigate prevalence, treatment, and control of hypercholesterolemia, included 11,680 US-born citizens, 2,752 foreign born citizens, and 2,554 non-citizens (Guadamuz et al. 2020). In that study, over half of non-citizens did not have health insurance (52.2); which was significantly more than US-born citizens (13.6%, p<0.001) (Guadamuz et al. 2020).

Non-citizens also had significantly higher prevalence of diabetes (15.7% vs. 12.8%, p<0.001) (Guadamuz et al. 2020). Treatment percentages were also significantly lower among non-citizens than US-born citizens with hypercholesteremia (16.4% vs 45.5%), hypertension (60.3% vs. 81.1%), and diabetes (51.2% vs. 69.5%) (p<0.001) (Guadamuz et al. 2020). Among noncitizens, those without a usual source of health care or health insurance had lower treatment percentages for hypercholesteremia (2.7% and 8.1%), hypertension (22.2% and 39.1%), and diabetes (15.5% and 28.6%) (Guadamuz et al. 2020). It is very important to understand that overall, environmental risk factors of the many pesticides on the global market are still poorly characterized across the literature.

### 1.3.2 Limitations and Strengths

Our research shows that NHANES respondents are exposed to multiple pesticides and pesticide types. Quantifying chemical mixtures across a population is complex and methodology for understanding these mixtures is still an emerging area of research. However, there is still plenty of research to be done in understanding chemical mixtures. Much of the research on chemical health outcomes focuses on one chemical at a time, including our study, but people are often exposed to more than one chemical, chemicals can interact with each other to create new chemicals and once chemicals are in the environment, they can also react with the ambient air or be degraded by the sun’s rays. All these changes to chemicals in relation to mixtures and being in the environment create nuanced exposures and further research is needed to understand how these mixtures may uniquely affect the human body.

Some pesticides which did not meet our inclusion criteria could have different exposure based on farmwork occupational status. Oxypyrimidine (7.88% vs. 13.76%, 0.033), desethyl hydroxy DEET (17.37% vs. 11.30%, p =0.015), and DEET (9.17% vs 6.25%, p = 0.036) were significantly different between farmworkers and non-farmworkers, respectively. However, all of these chemicals had detectability percentages below the cutoff for inclusion in our study. It is possible that by restricting the chemicals included we are missing some important differences in pesticide exposure between farmworkers and non-farmworkers. Studying exposures and effects of these less commonly detected pesticides could be an important area of investigation.

One of the major limitations of this project is that while NHANES is thorough, reliable, and valid study, it is still cross-sectional. This means the measurements within it are a single measurement in time and cannot be fully representative of chronic exposures or chronic symptomology due to exposures. Another limitation includes most farmworkers being recruited between 1999 and 2004 (N= 1,775, 69.6%), which is of importance since the recruitment and laboratory methods have been updated since 2003. Newer methods for quantifying chemicals from blood and urine samples are more sensitive and can detect lower quantities of chemicals. Additionally, farmworkers living without citizenship had significantly lower BMI as well, which may impact metabolism and accumulation of chemicals in the body.

An additional limitation of this study is that not every chemical is measured in every participant, and that not every assay is completed in each chemical. This limitation makes direct comparisons impossible and therefore our results are somewhat limited to group means. There are some known limitations to the ToxCast dataset such as interference of cytotoxicity. Non-specific cell stress can interfere with the frequency reading since the cell is overworking to re-gain homeostasis after chemical exposure. ToxCast assays are often assessing effects in a single tissue cell type, which may not accurately reflect chemical sensitivity across organ systems or within particularly susceptible individuals. Moreover, while ToxCast maintains a robust suite of assays measuring effects across a broad spectrum of potential toxic outcomes, not every chemical is tested for every assay and not all potential biological outcomes following chemical exposure are captured.

Other limitations inherent to interpreting bioactivity also exist. For starters, urine and serum concentrations reflect excreted or circulating concentrations, respectively, but may not be representative of concentrations in target organs like fat, liver, kidneys, or brain. This is important because many chemicals target specific organs (e.g., organochlorines targeting the central nervous system) or bioaccumulate in specific tissue types like lipids. There are also challenges to being able to relate metabolites to their parent compounds since some chemicals can have more than one parent compound (e.g. the pyrethroid metabolite 3-PBA). This can make ascertaining what active ingredient is bioactive in the human body difficult, and even if considering a limited number of chemicals, there is no way to calculate a direct contribution of each parent compound to a non-specific metabolite.

A strength of our study is that it is the first to provide a comprehensive quantification of all the pesticide exposure concentrations within the US population using NHANES from 1999 to 2014 and to then stratify these concentrations by social determinants of health with a focus on farmwork, fishing, and forestry work history and U.S. citizenship. By considering all the pesticides within NHANES and narrowing down to those with at least 50% detectability, we find that even within NHANES a small portion (15%) of these chemicals are detected in a majority of NHANES participants. ToxCast & NHANES are both validated, reliable study datasets created by the US government to assess chemical bioactivity and examine the health of people residing in the US. By integrating these two datasets, the results are more generalizable to the U.S. population. Additionally, this study is one of few to consider health disparities associated with occupation or citizenship and how they may affect pesticide exposure and potential resultant health effects. This project can inform evidence-based guidelines and policies that are focused on reducing pesticide exposure concentrations among people residing within the United States.

### 1.3.3 Future Directions

While NHANES quantifies many chemical biomarker concentrations for each study participant, these measures do not fully capture how many chemicals each person may be exposed to since every chemical is not tested for in every person. Moreover, toxicological research should continue to focus on novel methods for assessing toxicity of chemical mixtures and interactions to better understand population pesticide exposure and bioactivity of combined pesticide exposures in at-risk individuals. Currently, research looks at predominantly the active ingredients of pesticides, but inactive ingredients used to create pesticides may also influence human health, this is currently being missed in many toxicological studies. Future research can also include temporal data on pesticide exposure. Both NHANES and ToxCast include singular exposure time points in humans and *in vitro*, respectively. However, for many farmworkers, pesticide exposure is chronic and happens over multiple exposure incidents.

Expanding this research to disease biomarkers, symptoms, and diagnoses will also be an important future direction. This way we can better connect target families of ToxCast assays to health outcomes and then stratify findings by occupation and social determinants of health like income, gender, citizenship, and country of birth. In this same vein of understanding social determinant effects on health, more research on how these biomarker concentration distributions differ based on residing or working in a low versus high income country will be important because laws within a nation can alter the health and exposure for many.

## Data Availability

The NHANES and ToxCast datasets analysed during the current study are available from the CDC, https://wwwn.cdc.gov/nchs/nhanes/, and the EPA, https://www.epa.gov/chemical-research/exploring-toxcast-data.

https://www.epa.gov/chemical-research/exploring-toxcast-data

## Statements and Declarations

### Author Contributions

Justin Colacino and Chanese Forté contributed to the study conception and design. Material preparation and data collection were performed by Chanese Forté. Analysis was performed by Jess Millar and Chanese Forté. The first draft of the manuscript was written by Chanese Forté and all authors commented on subsequent versions of the manuscript. All authors read and approved the final manuscript.

