## Supplementary Tables for "Integrating NHANES and Toxicity Forecaster Data to Compare Pesticide Exposure and Bioactivity by Farmwork History and US Citizenship"

*Supplementary Table 1 Detectability of Chemicals by Work Category*

| Comment Code | Chemical Name | Non-Farmworkers Below LOD | Non-Farmworkers Above LOD | Non-Farmworkers % Above LOD | Farmworkers Below LOD | Farmworkers Above LOD | Farmworkers % Above LOD | *p*-value |
|---|---|---|---|---|---|---|---|---|
| LBXPDE | p,p'-DDE | 7 | 4455 | 99.84 | - | 274 | 100.00 | 1.000 |
| URX14D | 2,5-Dichlorophenol | 157 | 8513 | 98.19 | 7 | 278 | 97.54 | 0.369 |
| URXDCB | 2,4-Dichlorophenol | 1076 | 7594 | 87.59 | 42 | 243 | 85.26 | 0.237 |
| LBXTNA | Trans-nonachlor | 609 | 3828 | 86.27 | 25 | 245 | 90.74 | 0.035 |
| URXCPF | Chlorpyrifos | 903 | 5536 | 85.98 | 47 | 216 | 82.13 | 0.086 |
| URXCPM | 3,5,6-Trichloropyridinol | 903 | 5536 | 85.98 | 47 | 216 | 82.13 | 0.086 |
| URXCPO | Chlorpyrifos-oxon | 903 | 5536 | 85.98 | 47 | 216 | 82.13 | 0.086 |
| URXDEA | Deet Acid | 1327 | 5811 | 81.41 | 28 | 164 | 85.42 | 0.187 |
| URXOPM | 3-Phenoxybenzoic acid | 1537 | 6501 | 80.88 | 71 | 243 | 77.39 | 0.126 |
| LBXOXY | Oxychlordane | 1036 | 3134 | 75.16 | 37 | 219 | 85.55 | 8.87 e-05 |
| URXPAR | 4-Nitrophenol | 2049 | 5862 | 74.10 | 130 | 175 | 57.38 | 7.50 e-10 |
| LBXDIE | Dieldrin | 849 | 2144 | 71.63 | 51 | 144 | 73.85 | 0.566 |
| LBXBHC | Beta-hexachlorocyclohexane | 1350 | 3067 | 69.44 | 53 | 215 | 80.22 | 1.48 e-04 |
| URX24DCP | 2,4-Dichlorophenoxyacetic acid | 3713 | 5825 | 61.07 | 165 | 227 | 57.91 | 0.224 |
| LBXHPE | Heptachlor Epoxide | 1950 | 2171 | 52.68 | 107 | 153 | 58.85 | 0.055 |
| LBXPDT | p,p'-DDT | 2164 | 2047 | 48.61 | 109 | 155 | 58.71 | 1.49 e-03 |
| | *Below 48% Detectable* | | | | | | | |
| URXCCC | Cis Dichlorovnl-Dimeth Carboacid | 1667 | 1040 | 38.42 | 101 | 64 | 38.79 | 0.934 |
| LBXHCB | Hexachlorobenzene | 2703 | 1572 | 36.77 | 167 | 99 | 37.22 | 0.896 |
| URXALA | Alachlor mercapture | 613 | 334 | 35.27 | 31 | 17 | 35.42 | 1.000 |
| URXPCP | Pentachlorophenol | 1016 | 486 | 32.36 | 51 | 29 | 36.25 | 0.465 |
| URX1TB | 2,4,5-Trichlorophenol | 4781 | 2221 | 31.72 | 156 | 84 | 35.00 | 0.291 |
| LBXMIR | Mirex | 3107 | 1248 | 28.66 | 196 | 73 | 27.14 | 0.627 |
| URX3TB | 2,4,6-Trichlorophenol | 5003 | 1999 | 28.55 | 177 | 63 | 26.25 | 0.468 |
| URXMAL | Malathion Diacid | 2079 | 783 | 27.36 | 82 | 33 | 28.70 | 0.750 |

| | | | | | | | | |
|---|---|---|---|---|---|---|---|---|
| URXOPP | O-Phenyl Phenol | 5227 | 1775 | 25.35 | 168 | 72 | 30.00 | 0.114 |
| URXOXY | Oxypyrimidine | 4226 | 1121 | 20.97 | 121 | 26 | 17.69 | 0.410 |
| URXTCC | Desisopropyl Atrazine | 6348 | 1593 | 20.06 | 240 | 70 | 22.58 | 0.279 |
| URXDIZ | Oxypyrimidine | 2268 | 362 | 13.76 | 152 | 13 | 7.88 | 0.033 |
| URXDHD | Desethyl Hydroxy Deet | 6345 | 808 | 11.30 | 157 | 33 | 17.37 | 0.015 |
| URXETU | Ethylenethio urea | 4453 | 428 | 8.77 | 161 | 12 | 6.94 | 0.492 |
| URX4FP | Fluoro-Phenoxybenzoic acid | 7478 | 591 | 7.32 | 296 | 16 | 5.13 | 0.180 |
| URXDEE | DEET | 9264 | 618 | 6.25 | 327 | 33 | 9.17 | 0.036 |
| URXDPY | Diethylaminomethylpyrimidinol/One | 1616 | 106 | 6.16 | 109 | 6 | 5.22 | 0.841 |
| URXMET | Metolachlor Mercapturate | 1690 | 65 | 3.70 | 116 | 4 | 3.33 | 1.000 |
| URXCMH | Chloro-Hydro-Meth-Chromen-One/ Ol | 1647 | 62 | 3.63 | 114 | 3 | 2.56 | 0.796 |
| LBXODT | o,p'-DDT | 4029 | 139 | 3.33 | 251 | 11 | 4.20 | 0.478 |
| URXCBF | Carbofuranphenol | 4109 | 129 | 3.04 | 237 | 9 | 3.66 | 0.567 |
| URX25T | 2,4,5-Trichlorophenoxyacetic acid | 5926 | 165 | 2.71 | 310 | 6 | 1.90 | 0.476 |
| URXACE | Acetochlor Mercapturate | 1686 | 43 | 2.49 | 118 | 1 | 0.84 | 0.361 |
| URXAPE | Acephate | 4919 | 92 | 1.84 | 179 | 4 | 2.19 | 0.582 |
| LBXGHC | Gamma-hexachlorocyclohexane | 4240 | 70 | 1.62 | 258 | 8 | 3.01 | 0.135 |
| URXATZ | Atrazine mercapture | 4399 | 50 | 1.12 | 206 | 4 | 1.90 | 0.307 |
| URXCB3 | Deisopropyl Atrazine Mercapture | 4474 | 36 | 0.80 | 223 | 2 | 0.89 | 0.702 |
| URXDCZ | Diaminochloroatrazine | 1821 | 7 | 0.38 | 45 | 1 | 2.17 | 0.181 |
| URXPPX | 2-Isopropoxyphenol | 4163 | 16 | 0.38 | 237 | 4 | 1.66 | 0.021 |
| URXMMI | Methamidaphos | 5004 | 19 | 0.38 | 176 | 1 | 0.56 | 0.500 |
| URXMTO | Dimethoate | 5080 | 13 | 0.26 | 182 | - | 0.00 | 1.000 |
| URXPTU | Propylenethio urea | 5086 | 9 | 0.18 | 185 | - | 0.00 | 1.000 |
| LBXALD | Aldrin | 3063 | 5 | 0.16 | 196 | 1 | 0.51 | 0.312 |
| URXEMM | Ethametsulfuron Methyl | 4902 | 8 | 0.16 | 176 | - | 0.00 | 1.000 |
| URXNOS | Nicosulfuron | 4845 | 7 | 0.14 | 176 | - | 0.00 | 1.000 |
| URXDTZ | Desethyl Atrazine | 1753 | 2 | 0.11 | 44 | 1 | 2.22 | 0.073 |
| LBXEND | Endrin | 2913 | 3 | 0.10 | 191 | 1 | 0.52 | 0.225 |
| URXSIS | Desisopropyl Atrazine | 1722 | 1 | 0.06 | 44 | - | 0.00 | 1.000 |
| URXSIM | Desisopropyl Atrazine Mercapturate | 1752 | 1 | 0.06 | 45 | 1 | 2.17 | 0.051 |
| URXOMO | O-methoate | 5097 | 2 | 0.04 | 182 | - | 0.00 | 1.000 |

| | | | | | | | | |
|---|---|---|---|---|---|---|---|---|
| URXCHS | Chloro Sulfuron | 4725 | 1 | 0.02 | 169 | - | 0.00 | 1.000 |
| URXMTM | Metsulfuron Methyl | 4964 | 1 | 0.02 | 180 | - | 0.00 | 1.000 |
| URXSSF | Sulfosulfuron | 4999 | 1 | 0.02 | 181 | - | 0.00 | 1.000 |
| URXOXS | Oxasulfuron | 5013 | 1 | 0.02 | 181 | - | 0.00 | 1.000 |
| URXAAZ | Atrazine | 1828 | - | 0.00 | 46 | - | 0.00 | 1.000 |
| URXBSM | Bensulfuron Methyl | 4974 | - | 0.00 | 181 | - | 0.00 | 1.000 |
| URXFRM | Foramsulfuron | 4780 | - | 0.00 | 174 | - | 0.00 | 1.000 |
| URXHLS | Halosulfuron | 4903 | - | 0.00 | 181 | - | 0.00 | 1.000 |
| URXMSM | Mesosulfuron Methyl | 5020 | - | 0.00 | 181 | - | 0.00 | 1.000 |
| URXPIM | Primisulfuron Methyl | 4735 | - | 0.00 | 174 | - | 0.00 | 1.000 |
| URXPRO | Prosulfuron | 4843 | - | 0.00 | 176 | - | 0.00 | 1.000 |
| URXRIM | Rimsulfuron | 4938 | - | 0.00 | 175 | - | 0.00 | 1.000 |
| URXSMM | Sulfometuron Methyl | 4759 | - | 0.00 | 169 | - | 0.00 | 1.000 |
| URXTHF | Thifensulfuron Methyl | 4974 | - | 0.00 | 179 | - | 0.00 | 1.000 |
| URXTRA | Triasulfuron | 4872 | - | 0.00 | 178 | - | 0.00 | 1.000 |
| URXTRN | Triflusulfuron Methyl | 4967 | - | 0.00 | 183 | - | 0.00 | 1.000 |

Data obtained from NHANES, an abbreviation for the National Health and Nutrition Examination Survey, a cross-sectional study of people residing in the United States and maintained by the Centers for Disease Control and Prevention. The above chemicals are included based on respondent (unique SEQN) who also had occupation data and laboratory results data collected.

*Supplementary Table 2 Non-parametric Wilcoxon-Mann-Whitney U Table*

| Chemical/ Comparison | Weights | ESS | *N* | *t* | Df | ROC | *p* | *p* FDR | |
|---|---|---|---|---|---|---|---|---|---|
| **ß-Hexachlorocyclohexane A** | | | | | | | | | |
| Farmworker vs Non | All | 261 | 4685 | 1.9292 | 43 | 0.5605 | 0.0603 | 0.1086 | |
| Non-US Citizen vs US Citizen | All | 620 | 6129 | 13.9573 | 43 | 0.7152 | 1.4276e-17 | 5.1394e-16 | * |
| **Dieldrin** | | | | | | | | | |
| Farmworker vs Non | All | 194 | 3188 | 0.8164 | 29 | 0.5267 | 0.4209 | 0.4736 | |
| Non-US Citizen vs US Citizen | All | 420 | 4109 | -3.0742 | 29 | 0.4098 | 4.5658e-03 | 0.0110 | * |
| **Heptachlor Epoxide** | | | | | | | | | |
| Farmworker vs Non | All | 253 | 4381 | 1.3660 | 43 | 0.5371 | 0.1790 | 0.2482 | |
| Non-US Citizen vs US Citizen | All | 586 | 5799 | -3.3394 | 43 | 0.4812 | 1.7426e-03 | 5.2279e-03 | * |
| **p,p'-DDE** | | | | | | | | | |
| Farmworker vs Non | All | 267 | 4736 | 1.5872 | 43 | 0.5605 | 0.1198 | 0.1960 | |
| Non-US Citizen vs US Citizen | All | 625 | 6202 | 9.4831 | 43 | 0.7194 | 4.2143e-12 | 7.5857e-11 | * |
| **p,p'-DDT** | | | | | | | | | |
| Farmworker vs Non | All | 255 | 4475 | -0.5800 | 43 | 0.5292 | 0.5650 | 0.6163 | |
| Non-US Citizen vs US Citizen | All | 595 | 5937 | 6.5620 | 43 | 0.6902 | 5.5614e-08 | 6.6736e-07 | * |
| **2,4-Dichlorophenoxyacetic Acid** | | | | | | | | | |
| Farmworker vs Non | All | 355 | 9926 | 2.2979 | 91 | 0.5230 | 0.0239 | 0.0452 | * |
| Non-US Citizen vs US Citizen | All | 1549 | 15120 | -3.2651 | 91 | 0.4671 | 1.5428e-03 | 5.0491e-03 | * |
| **DEET Acid** | | | | | | | | | |
| Farmworker vs Non | All | 158 | 7328 | 1.7931 | 63 | 0.5634 | 0.0778 | 0.1333 | |
| Non-US Citizen vs US Citizen | All | 1018 | 10343 | -3.1475 | 63 | 0.4368 | 2.5154e-03 | 6.9657e-03 | * |
| **3-Phenoxybenzoic Acid** | | | | | | | | | |
| Farmworker vs Non | All | 267 | 8349 | -0.4472 | 76 | 0.4585 | 0.6560 | 0.6946 | |
| Non-US Citizen vs US Citizen | All | 1326 | 12784 | -1.1166 | 76 | 0.4800 | 0.2677 | 0.3323 | |
| **4-Nitrophenol** | | | | | | | | | |
| Farmworker vs Non | All | 260 | 8213 | -4.3256 | 76 | 0.4141 | 4.5680e-05 | 2.7408e-04 | * |
| Non-US Citizen vs US Citizen | All | 1308 | 12614 | 2.4865 | 76 | 0.5266 | 0.0151 | 0.0302 | * |
| **3,5,6-Trichloropyridinol** | | | | | | | | | |
| Farmworker vs Non | All | 219 | 6699 | 0.2903 | 59 | 0.5093 | 0.7726 | 0.7947 | |
| Non-US Citizen vs US Citizen | All | 1082 | 10368 | -3.0750 | 59 | 0.4608 | 3.1867e-03 | 8.1943e-03 | * |

| | | | | | | | | | |
|---|---|---|---|---|---|---|---|---|---|
| **2,5-Dichlorophenol** | | | | | | | | | |
| Farmworker vs Non | All | 256 | 8953 | -0.8212 | 78 | 0.4957 | 0.4140 | 0.4736 | |
| Non-US Citizen vs US Citizen | All | 1192 | 12885 | 5.4372 | 78 | 0.5827 | 6.0034e-07 | 5.4030e-06 | * |
| Farmworker vs Non | Sub A | 45 | 1712 | -1.2505 | 16 | 0.4403 | 0.2291 | 0.2946 | |
| Non-US Citizen vs US Citizen | Sub A | 264 | 2474 | 4.0748 | 16 | 0.5858 | 8.8191e-04 | 3.5276e-03 | * |
| Farmworker vs Non | Sub B | 116 | 5550 | -0.2446 | 46 | 0.5093 | 0.8079 | 0.8079 | |
| Non-US Citizen vs US Citizen | Sub B | 699 | 7890 | 3.7415 | 46 | 0.5880 | 5.0636e-04 | 2.5898e-03 | * |
| Farmworker vs Non | Sub C | 95 | 1691 | -1.4778 | 14 | 0.4550 | 0.1616 | 0.2424 | |
| Non-US Citizen vs US Citizen | Sub C | 242 | 2521 | 3.0665 | 14 | 0.5820 | 8.3706e-03 | 0.0188 | * |
| **2,4-Dichlorophenol** | | | | | | | | | |
| Farmworker vs Non | All | 256 | 8953 | -1.0113 | 78 | 0.5106 | 0.3150 | 0.3780 | |
| Non-US Citizen vs US Citizen | All | 1192 | 12885 | 4.8235 | 78 | 0.5753 | 6.8516e-06 | 4.9332e-05 | * |
| Farmworker vs Non | Sub A | 45 | 1712 | -1.5022 | 16 | 0.4997 | 0.1525 | 0.2387 | |
| Non-US Citizen vs US Citizen | Sub A | 264 | 24747 | 3.8880 | 16 | 0.5861 | 1.3064e-03 | 4.7030e-03 | * |
| Farmworker vs Non | Sub B | 116 | 5550 | 1.3228 | 46 | 0.5528 | 0.1924 | 0.2566 | |
| Non-US Citizen vs US Citizen | Sub B | 699 | 7890 | 3.6993 | 46 | 0.5860 | 5.7552e-04 | 2.5898e-03 | * |
| Farmworker vs Non | Sub C | 95 | 1691 | -2.8901 | 14 | 0.4311 | 0.0119 | 0.0251 | * |
| Non-US Citizen vs US Citizen | Sub C | 242 | 2521 | 1.4139 | 14 | 0.5440 | 0.1793 | 0.2482 | |

Wilcoxon-Mann-Whitney U ranks molarity of chemicals to inquire if two groups are statistically different from one another. Chemicals are considered here as the log molarity of the chemical ($\mu$mol/$L$) for blood measurements and log ratio of the molarity of the chemical with the log molarity of creatine ($\mu$mo/l$L$) for urinary measurements.

*Supplementary Table 3 Unadjusted Logistic Regression Table*

| Chemical & Comparison | Weights | ESS | N | OR (95% CI) | | t | ROC | *p* | *p* FDR | |
|---|---|---|---|---|---|---|---|---|---|---|
| **ß-Hexachlorocyclohexane A** | | | | | | | | | | |
| Farmworker vs Non | All | 261 | 4685 | 1.24 | (0.91, 1.70) | 1.40 | 0.51 | 0.17 | 0.27 | |
| Non-US Citizen vs US Citizen | All | 342 | 6129 | 3.27 | (2.66, 4.03) | 11.58 | 0.54 | 8.49e-15 | 2.21e-13 | * |
| **Dieldrin** | | | | | | | | | | |
| Farmworker vs Non | All | 194 | 3188 | - | - | - | - | - | - | |
| Non-US Citizen vs US Citizen | All | 250 | 4109 | - | - | - | - | - | - | |
| **Heptachlor Epoxide** | | | | | | | | | | |
| Farmworker vs Non | All | 253 | 4381 | - | - | - | - | - | - | |
| Non-US Citizen vs US Citizen | All | 334 | 5799 | - | - | - | - | - | - | |
| **p,p'-DDE** | | | | | | | | | | |
| Farmworker vs Non | All | 267 | 4736 | 1.23 | (0.81, 1.85) | 0.99 | 0.51 | 0.33 | 0.47 | |
| Non-US Citizen vs US Citizen | All | 350 | 6202 | 4.31 | (2.63, 7.05) | 5.98 | 0.53 | 3.96e-07 | 2.06e-06 | * |
| **p,p'-DDT** | | | | | | | | | | |
| Farmworker vs Non | All | 255 | 4475 | 1.55 | (0.52, 4.65) | 0.81 | 0.51 | 0.42 | 0.54 | |
| Non-US Citizen vs US Citizen | All | 336 | 5937 | 31.16 | (13.38, 72.55) | 8.21 | 0.82 | 2.42e-10 | 3.15e-09 | * |
| **2,4-Dichlorophenoxyacetic Acid** | | | | | | | | | | |
| Farmworker vs Non | All | 330 | 9460 | 4.14 | (2.60, 6.60) | 6.04 | 0.54 | 3.27e-08 | 2.12e-07 | * |
| Non-US Citizen vs US Citizen | All | 527 | 14636 | 0.93 | (0.66, 1.30) | -0.45 | 0.50 | 0.65 | 0.77 | |
| **DEET Acid** | | | | | | | | | | |
| Farmworker vs Non | All | 158 | 7330 | 2.39 | (1.01, 5.63) | 2.03 | 0.51 | 0.05 | 0.10 | |
| Non-US Citizen vs US Citizen | All | 220 | 10346 | 0.73 | (0.50, 1.06) | -1.68 | 0.51 | 0.10 | 0.18 | |
| **3-Phenoxybenzoic Acid** | | | | | | | | | | |
| Farmworker vs Non | All | 241 | 7864 | - | - | - | - | - | - | |
| Non-US Citizen vs US Citizen | All | 395 | 12279 | - | - | - | - | - | - | |
| **4-Nitrophenol** | | | | | | | | | | |
| Farmworker vs Non | All | 234 | 7738 | 0.63 | (0.43, 0.93) | -2.34 | 0.51 | 0.02 | 0.06 | |
| Non-US Citizen vs US Citizen | All | 386 | 12120 | 1.19 | (1.00, 1.40) | 2.04 | 0.51 | 0.04 | 0.10 | |
| **3,5,6-Trichloropyridinol** | | | | | | | | | | |
| Farmworker vs Non | All | 193 | 6220 | - | - | - | - | - | - | |
| Non-US Citizen vs US Citizen | All | 329 | 9870 | - | - | - | - | - | - | |

| | | | | | | | | | | |
|---|---|---|---|---|---|---|---|---|---|---|
| **2,5-Dichlorophenol** | | | | | | | | | | |
| Farmworker vs Non | All | 256 | 8955 | 1.04 | (0.75, 1.44) | 0.24 | 0.50 | 0.81 | 0.90 | |
| Non-US Citizen vs US Citizen | All | 368 | 12890 | 2.02 | (1.61, 2.54) | 6.15 | 0.53 | 3.11e-08 | 2.12e-07 | * |
| Farmworker vs Non | Sub A | 45 | 1713 | 0.48 | (0.19, 1.19) | -1.71 | 0.51 | 0.11 | 0.18 | |
| Non-US Citizen vs US Citizen | Sub A | 65 | 2476 | 2.26 | (1.45, 3.52) | 3.89 | 0.54 | 1.30e-03 | 3.76e-03 | * |
| Farmworker vs Non | Sub B | 116 | 5550 | 1.29 | (0.75, 2.21) | 0.93 | 0.50 | 0.36 | 0.49 | |
| Non-US Citizen vs US Citizen | Sub B | 163 | 7890 | 2.14 | (1.60, 2.87) | 5.28 | 0.53 | 3.39e-06 | 1.47e-05 | * |
| Farmworker vs Non | Sub C | 95 | 1692 | 0.84 | (0.53, 1.33) | -0.81 | 0.50 | 0.43 | 0.54 | |
| Non-US Citizen vs US Citizen | Sub C | 141 | 2524 | 1.76 | (1.00, 3.10) | 2.13 | 0.52 | 0.05 | 0.10 | |
| **2,4-Dichlorophenol** | | | | | | | | | | |
| Farmworker vs Non | All | 256 | 8955 | 0.97 | (0.55, 1.70) | -0.12 | 0.50 | 0.90 | 0.90 | |
| Non-US Citizen vs US Citizen | All | 368 | 12890 | 2.63 | (1.52, 4.54) | 3.50 | 0.55 | 7.63e-04 | 2.48e-03 | * |
| Farmworker vs Non | Sub A | 45 | 1713 | - | - | - | - | - | - | |
| Non-US Citizen vs US Citizen | Sub A | 65 | 2476 | - | - | - | - | - | - | |
| Farmworker vs Non | Sub B | 116 | 5550 | 0.92 | (0.28, 3.06) | -0.13 | 0.50 | 0.89 | 0.90 | |
| Non-US Citizen vs US Citizen | Sub B | 163 | 7890 | 3.76 | (1.94, 7.30) | 4.02 | 0.58 | 2.12e-04 | 7.87e-04 | * |
| Farmworker vs Non | Sub C | 95 | 1692 | 1.06 | (0.47, 2.36) | 0.14 | 0.50 | 0.89 | 0.90 | |
| Non-US Citizen vs US Citizen | Sub C | 141 | 2524 | 1.83 | (0.69, 4.84) | 1.34 | 0.53 | 0.20 | 0.31 | |

Logistic model testing the odds of having a bioactive measurement of a given chemical. A bioactive measurement is defined as a positive hit call.

*Supplementary Table 4 Adjusted Logistic Regression Table*

| Chemical & Comparison | Weights | ESS | N | OR (95% CI) | | t | ROC | *p* | *p* FDR | |
|---|---|---|---|---|---|---|---|---|---|---|
| **ß-Hexachlorocyclohexane A** | | | | | | | | | | |
| Farmworker vs Non | All | 253 | 4570 | 0.99 | (0.63, 1.55) | -0.03 | 0.88 | 0.97 | 0.97 | |
| Non-US Citizen vs US Citizen | All | 332 | 5982 | 8.50 | (5.45, 13.24) | 9.84 | 0.90 | 4.75e-11 | 1.23e-09 | * |
| **Dieldrin** | | | | | | | | | | |
| Farmworker vs Non | All | 187 | 3092 | - | - | - | - | - | - | |
| Non-US Citizen vs US Citizen | All | 241 | 3988 | - | - | - | - | - | - | |
| **Heptachlor Epoxide** | | | | | | | | | | |
| Farmworker vs Non | All | 253 | 4272 | - | - | - | - | - | - | |
| Non-US Citizen vs US Citizen | All | 334 | 5659 | - | - | - | - | - | - | |
| **p,p'-DDE** | | | | | | | | | | |
| Farmworker vs Non | All | 267 | 4620 | 0.93 | (0.63, 1.36) | -0.39 | 0.79 | 0.70 | 0.89 | |
| Non-US Citizen vs US Citizen | All | 350 | 6054 | 2.98 | (1.69, 5.29) | 3.90 | 0.81 | 4.80e-04 | 3.12e-03 | * |
| **p,p'-DDT** | | | | | | | | | | |
| Farmworker vs Non | All | 247 | 4362 | 1.49 | (0.51, 4.35) | 0.75 | 0.90 | 0.46 | 0.76 | |
| Non-US Citizen vs US Citizen | All | 326 | 5792 | 10.77 | (3.63, 31.95) | 4.46 | 0.92 | 1.00e-04 | 8.70e-04 | * |
| **2,4-Dichlorophenoxyacetic Acid** | | | | | | | | | | |
| Farmworker vs Non | All | 321 | 9326 | 4.33 | (2.73, 6.87) | 6.31 | 0.73 | 1.55e-08 | 2.01e-07 | * |
| Non-US Citizen vs US Citizen | All | 494 | 13941 | 1.05 | (0.69, 1.61) | 0.23 | 0.70 | 0.82 | 0.93 | |
| **DEET Acid** | | | | | | | | | | |
| Farmworker vs Non | All | 157 | 7253 | 2.23 | (0.97, 5.13) | 1.94 | 0.65 | 0.06 | 0.22 | |
| Non-US Citizen vs US Citizen | All | 212 | 9928 | 0.89 | (0.62, 1.27) | -0.66 | 0.64 | 0.51 | 0.76 | |
| **3-Phenoxybenzoic Acid** | | | | | | | | | | |
| Farmworker vs Non | All | 234 | 7763 | - | - | - | - | - | - | |
| Non-US Citizen vs US Citizen | All | 371 | 11679 | - | - | - | - | - | - | |
| **4-Nitrophenol** | | | | | | | | | | |
| Farmworker vs Non | All | 228 | 7639 | 0.57 | (0.38, 0.87) | -2.69 | 0.76 | 9.15e-03 | 0.04 | * |
| Non-US Citizen vs US Citizen | All | 362 | 11523 | 1.17 | (0.95, 1.44) | 1.53 | 0.75 | 0.13 | 0.37 | |
| **3,5,6-Trichloropyridinol** | | | | | | | | | | |
| Farmworker vs Non | All | 187 | 6139 | - | - | - | - | - | - | |
| Non-US Citizen vs US Citizen | All | 307 | 9388 | - | - | - | - | - | - | |

| | | | | | | | | | |
|---|---|---|---|---|---|---|---|---|---|
| **2,5-Dichlorophenol** | | | | | | | | | |
| Farmworker vs Non | All | 251 | 8832 | 1.01 | (0.75, 1.35) | 0.06 | 0.73 | 0.95 | 0.97 |
| Non-US Citizen vs US Citizen | All | 351 | 12400 | 1.35 | (1.02, 1.79) | 2.16 | 0.73 | 0.03 | 0.15 |
| Farmworker vs Non | Sub A | 43 | 1681 | 0.63 | (0.23, 1.73) | -1.27 | 0.71 | 0.27 | 0.54 |
| Non-US Citizen vs US Citizen | Sub A | 61 | 2377 | 1.73 | (0.88, 3.43) | 2.25 | 0.71 | 0.09 | 0.29 |
| Farmworker vs Non | Sub B | 115 | 5490 | 1.04 | (0.64, 1.72) | 0.18 | 0.73 | 0.86 | 0.93 |
| Non-US Citizen vs US Citizen | Sub B | 157 | 7607 | 1.24 | (0.87, 1.79) | 1.23 | 0.74 | 0.23 | 0.54 |
| Farmworker vs Non | Sub C | 93 | 1657 | 1.09 | (0.45, 2.63) | 0.41 | 0.74 | 0.72 | 0.89 |
| Non-US Citizen vs US Citizen | Sub C | 133 | 2416 | 1.30 | (0.31, 5.57) | 0.79 | 0.73 | 0.51 | 0.76 |
| **2,4-Dichlorophenol** | | | | | | | | | |
| Farmworker vs Non | All | 251 | 8832 | 0.86 | (0.48, 1.54) | -0.51 | 0.79 | 0.61 | 0.83 |
| Non-US Citizen vs US Citizen | All | 351 | 12400 | 1.51 | (0.72, 3.17) | 1.11 | 0.78 | 0.27 | 0.54 |
| Farmworker vs Non | Sub A | 43 | 1681 | - | - | - | - | - | - |
| Non-US Citizen vs US Citizen | Sub A | 61 | 2377 | - | - | - | - | - | - |
| Farmworker vs Non | Sub B | 115 | 5490 | 0.69 | (0.21, 2.26) | -0.64 | 0.85 | 0.53 | 0.76 |
| Non-US Citizen vs US Citizen | Sub B | 157 | 7607 | 2.07 | (0.77, 5.56) | 1.50 | 0.84 | 0.14 | 0.37 |
| Farmworker vs Non | Sub C | 93 | 1657 | 1.54 | (0.26, 8.93) | 1.05 | 0.80 | 0.40 | 0.75 |
| Non-US Citizen vs US Citizen | Sub C | 133 | 2416 | 0.81 | (0.05, 13.99) | -0.32 | 0.77 | 0.78 | 0.92 |

Logistic model testing the odds of having a bioactive measurement of a given chemical. Adjusted model includes adjustments for BMI, age, gender, education, and ethnicity. Log molarity of creatine is also added for urinary measurements.

*Supplementary Table 5 Adjusted Linear Regression Table*

| Chemical & Comparison | Weights | ESS | N | ß (95% CI) | | t | $R^2$ | *p* | *p* FDR | |
|---|---|---|---|---|---|---|---|---|---|---|
| **ß-Hexachlorocyclohexane A** | | | | | | | | | | |
| Farmworker vs Non | All | 253 | 4570 | -0.02 | (-0.31, 0.28) | -0.11 | 0.39 | 0.91 | 0.95 | |
| Non-US Citizen vs US Citizen | All | 332 | 5982 | 2.99 | (1.57, 5.68) | 3.47 | 0.42 | 1.56e-03 | 0.01 | * |
| **Dieldrin** | | | | | | | | | | |
| Farmworker vs Non | All | 187 | 3092 | -0.07 | (-0.45, 0.30) | -0.40 | 0.20 | 0.69 | 0.89 | |
| Non-US Citizen vs US Citizen | All | 241 | 3988 | 0.94 | (0.78, 1.13) | -0.70 | 0.19 | 0.49 | 0.80 | |
| **Heptachlor Epoxide** | | | | | | | | | | |
| Farmworker vs Non | All | 245 | 4272 | 0.03 | (-0.10,0.16) | 0.47 | 0.26 | 0.64 | 0.86 | |
| Non-US Citizen vs US Citizen | All | 323 | 5659 | 0.91 | (0.77, 1.06) | -1.25 | 0.27 | 0.22 | 0.49 | |
| **p,p'-DDE** | | | | | | | | | | |
| Farmworker vs Non | All | 259 | 4620 | 0.28 | (-0.03, 0.58) | 1.84 | 0.36 | 0.07 | 0.34 | |
| Non-US Citizen vs US Citizen | All | 340 | 6054 | 2.51 | (1.91, 3.30) | 6.89 | 0.41 | 1.02e-07 | 1.83e-06 | * |
| **p,p'-DDT** | | | | | | | | | | |
| Farmworker vs Non | All | 247 | 4362 | 0.17 | (-0.22, 0.56) | 0.90 | 0.27 | 0.38 | 0.64 | |
| Non-US Citizen vs US Citizen | All | 326 | 5792 | 4.18 | (3.13, 5.60) | 10.03 | 0.35 | 2.98e-11 | 1.07e-09 | * |
| **2,4-Dichlorophenoxyacetic Acid** | | | | | | | | | | |
| Farmworker vs Non | All | 321 | 9326 | 0.48 | (-0.26, 1.23) | 1.29 | 0.29 | 0.20 | 0.48 | |
| Non-US Citizen vs US Citizen | All | 494 | 13941 | 0.94 | (0.75, 1.18) | -0.57 | 0.24 | 0.57 | 0.85 | |
| **DEET Acid** | | | | | | | | | | |
| Farmworker vs Non | All | 157 | 7253 | -0.03 | (-2.38, 1.77) | -0.30 | 0.46 | 0.77 | 0.92 | |
| Non-US Citizen vs US Citizen | All | 212 | 9928 | 0.91 | (0.27, 3.08) | -0.15 | 0.42 | 0.88 | 0.95 | |
| **3-Phenoxybenzoic Acid** | | | | | | | | | | |
| Farmworker vs Non | All | 234 | 7763 | -0.15 | (-0.69, 0.40) | -0.54 | 0.15 | 0.59 | 0.85 | |
| Non-US Citizen vs US Citizen | All | 371 | 11679 | 0.78 | (0.58, 1.06) | -1.61 | 0.14 | 0.11 | 0.41 | |
| **4-Nitrophenol** | | | | | | | | | | |
| Farmworker vs Non | All | 228 | 7639 | -0.55 | (-0.76, -0.35) | -5.35 | 0.12 | 1.30e-06 | 1.55e-05 | * |
| Non-US Citizen vs US Citizen | All | 362 | 11523 | 1.05 | (0.91, 1.21) | 0.66 | 0.11 | 0.51 | 0.80 | |
| **3,5,6-Trichloropyridinol** | | | | | | | | | | |
| Farmworker vs Non | All | 187 | 6139 | -0.11 | (-0.34, 0.13) | -0.92 | 0.33 | 0.36 | 0.64 | |
| Non-US Citizen vs US Citizen | All | 307 | 9388 | 0.91 | (0.74, 1.12) | -0.94 | 0.33 | 0.35 | 0.64 | |

| | | | | | | | | | |
|---|---|---|---|---|---|---|---|---|---|
| **2,5-Dichlorophenol** | | | | | | | | | |
| Farmworker vs Non | All | 251 | 8832 | -0.35 | (-0.69, -0.01) | -2.05 | 0.16 | 0.04 | 0.27 |
| Non-US Citizen vs US Citizen | All | 351 | 12400 | 1.52 | (0.91, 2.53) | 1.63 | 0.16 | 0.11 | 0.41 |
| Farmworker vs Non | Sub A | 43 | 1681 | -0.76 | (-2.07, 0.54) | -1.62 | 0.19 | 0.18 | 0.48 |
| Non-US Citizen vs US Citizen | Sub A | 61 | 2377 | 1.11 | (0.37, 3.34) | 0.25 | 0.16 | 0.81 | 0.94 |
| Farmworker vs Non | Sub B | 115 | 5494 | -0.05 | (-0.55, 0.45) | -0.20 | 0.20 | 0.85 | 0.95 |
| Non-US Citizen vs US Citizen | Sub B | 157 | 7607 | 1.71 | (0.82, 3.56) | 1.48 | 0.19 | 0.15 | 0.48 |
| Farmworker vs Non | Sub C | 93 | 1657 | -0.29 | (-1.26, 0.69) | -1.26 | 0.20 | 0.33 | 0.64 |
| Non-US Citizen vs US Citizen | Sub C | 133 | 2416 | 1.25 | (0.23, 6.87) | 0.57 | 0.17 | 0.63 | 0.86 |
| **2,4-Dichlorophenol** | | | | | | | | | |
| Farmworker vs Non | All | 251 | 8832 | -0.42 | (-0.72, -0.11) | -2.72 | 0.17 | 8.40e-03 | 0.06 |
| Non-US Citizen vs US Citizen | All | 351 | 12400 | 1.28 | (0.85, 1.93) | 1.21 | 0.16 | 0.23 | 0.49 |
| Farmworker vs Non | Sub A | 43 | 1681 | -0.75 | (-1.56, 0.07) | -2.54 | 0.20 | 0.06 | 0.33 |
| Non-US Citizen vs US Citizen | Sub A | 61 | 2377 | 1.03 | (0.40, 2.67) | 0.08 | 0.17 | 0.94 | 0.95 |
| Farmworker vs Non | Sub B | 115 | 5494 | -0.07 | (-0.52, 0.38) | -0.32 | 0.21 | 0.75 | 0.92 |
| Non-US Citizen vs US Citizen | Sub B | 157 | 7607 | 1.46 | (0.82, 2.59) | 1.33 | 0.20 | 0.19 | 0.48 |
| Farmworker vs Non | Sub C | 93 | 1657 | -0.45 | (-1.35, 0.45) | -2.17 | 0.18 | 0.16 | 0.48 |
| Non-US Citizen vs US Citizen | Sub C | 133 | 2416 | 1.02 | (0.22, 4.72) | 0.07 | 0.16 | 0.95 | 0.95 |

Linear model testing the unit change in molarity of a given chemical. Adjusted model includes adjustments for BMI, age, gender, education, and ethnicity. Log molarity of creatine is also added for urinary measurements.

*Supplementary Table 6 Toxcast Assay Intended Target Family Frequencies, by Pesticide*

| Intended Target Family | 2,4-D | 4-Nitrophenol | ß-HCH | p,p'-DDE | p,p'-DDT | Total |
|---|---|---|---|---|---|---|
| cell cycle | 7 | 28 | 6 | 74 | 120 | 235 |
| nuclear receptor | 8 | 4 | 12 | 102 | 58 | 184 |
| DNA binding | 2 | 0 | 3 | 64 | 27 | 96 |
| cytokine | 0 | 0 | 0 | 29 | 33 | 62 |
| cell adhesion molecules | 0 | 0 | 0 | 13 | 14 | 27 |
| cell morphology | 2 | 2 | 1 | 6 | 8 | 19 |
| GPCR | 1 | 0 | 0 | 4 | 4 | 9 |
| protease | 0 | 1 | 0 | 5 | 3 | 9 |
| histones | 0 | 0 | 0 | 2 | 2 | 4 |
| hydrolase | 0 | 2 | 1 | 0 | 1 | 4 |
| malformation | 1 | 1 | 0 | 1 | 1 | 4 |
| oxidoreductase | 1 | 3 | 0 | 0 | 0 | 4 |
| protease inhibitor | 0 | 0 | 0 | 2 | 2 | 4 |
| steroid hormone | 3 | 1 | 0 | 0 | 0 | 4 |
| transporter | 1 | 0 | 0 | 1 | 1 | 3 |
| growth factor | 0 | 0 | 0 | 1 | 1 | 2 |
| kinase | 0 | 0 | 0 | 1 | 1 | 2 |
| phosphatase | 1 | 1 | 0 | 0 | 1 | 3 |
| CYP | 0 | 0 | 1 | 0 | 0 | 1 |
| misc protein | 0 | 0 | 0 | 0 | 1 | 1 |
| *Total* | *27* | *43* | *24* | *305* | *278* | *677* |

Focusing on bioactive chemicals only, frequencies of intended target family for each chemical is presented here. Intended target family describes the category of the assay cellular effect. Bioactive is described as having a positive hit-call for the assay.
